# Identification of moderate effect size genes in autism spectrum disorder through a novel gene pairing approach

**DOI:** 10.1101/2024.04.03.24305278

**Authors:** Madison Caballero, F Kyle Satterstrom, Joseph D. Buxbaum, Behrang Mahjani

## Abstract

Autism Spectrum Disorder (ASD) arises from complex genetic and environmental factors, with inherited genetic variation playing a substantial role. This study introduces a novel approach to uncover moderate effect size (MES) genes in ASD, which individually do not meet the ASD liability threshold but collectively contribute when paired with specific other MES genes. Analyzing 10,795 families from the SPARK dataset, we identified 97 MES genes forming 50 significant gene pairs, demonstrating a substantial association with ASD when considered in tandem, but not individually. Our method leverages familial inheritance patterns and statistical analyses, refined by comparisons against control cohorts, to elucidate these gene pairs’ contribution to ASD liability. Furthermore, expression profile analyses of these genes in brain tissues underscore their relevance to ASD pathology. This study underscores the complexity of ASD’s genetic landscape, suggesting that gene combinations, beyond high impact single-gene mutations, significantly contribute to the disorder’s etiology and heterogeneity. Our findings pave the way for new avenues in understanding ASD’s genetic underpinnings and developing targeted therapeutic strategies.

## Introduction

Autism Spectrum Disorder (ASD) is a complex neurodevelopmental disorder characterized by challenges in social interaction and communication, as well as a tendency toward restricted and repetitive behaviors. Extensive research has underscored the complex interplay of genetic and environmental factors contributing to its risk ^1–6^. Diverse environmental influences, including pregnancy complications, parental age, pollutants, and socioeconomic status, contribute to the overall risk of ASD ^7^. Nonetheless, there is consensus that genetic factors exert greater influence on ASD’s etiology, underscored by its substantial heritability exceeding 50% ^2,8^. Large-scale genomic studies have further revealed that ASD is a polygenic disorder influenced by a broad spectrum of genetic variants rather than a single gene mutation ^9^. These variants, ranging in effect sizes, collectively shape the likelihood and clinical manifestations of ASD. The threshold liability model for ASD, defined by an individual’s unique combination of genetic and environmental factors, has become the predominant way to understand and evaluate the complex interplay of influences contributing to ASD risk and manifestation. Thus, the likelihood of developing ASD is dependent on whether the summation of these factors crosses the threshold for an ASD phenotype.

Genetic factors contributing to ASD liability range in rarity and effect size. Inherited common variants (those with a population frequency of at least 1%) explain approximately 50% of individual ASD liability ^1,2,6^. Though common variants play a substantial role when considered together, an individual common variant provides little contribution to individual ASD liability and often offers little biological insight. The pressure of negative selection forces variants that individually confer substantial heritability for ASD to be rare in the population; thus, studies of rare variants – ranging from single nucleotide variants to large structural changes – have been undertaken to help identify risk genes and developmental pathways. In recent years, research efforts have focused on identifying *de novo* mutations in particular. *De novo* loss-of-function variants are rare in the general population, particularly in genes which are haploinsufficient or which exhibit evolutionary constraint. Genes in which such variants are overrepresented in individuals with ASD can be identified as ASD risk genes, and the variants themselves can confer extreme phenotypes that provide meaningful biological insight ^3,4^. However, *de novo* mutations only account for approximately 5% of ASD cases ^2^, and their sporadic origin renders them less representative of ASD’s overall heritability.

While there are an estimated 1100 genes implicated in ASD ^10^, only a few hundred have been confidently identified ^3,4,6,11^. Rare variant analyses aggregate variants across a gene and employ advanced statistical models such as the Transmission and *De Novo* Association (TADA) test ^12^. TADA increases statistical power by combining *de novo* mutations and case-control inherited variants while incorporating per-gene mutation rates and gene constraint scores like the loss-of-function tolerance ^13,14^. Driven by variants of large effect, TADA has significantly improved the identification of genes associated with ASD ^4^. A notable set of genes, namely *SCN2A, SHANK3, CHD8, ADNP*, and *SYNGAP1*, demonstrate especially compelling evidence, as deleterious variants within these genes are effectively absent in control groups. However, this approach loses power when considering genes where deleterious variants have a smaller effect size, as these variants are more frequently found in non-ASD controls. As a result, there is a gap in our understanding of genes where variants make moderate contributions to ASD liability.

Here we propose and implement a novel method for identifying moderate effect size (MES) genes. We define MES genes as genes that individually do not surpass the ASD liability threshold when disrupted and therefore have no significant association when tested in isolation (**Fig 1A**). However, when deleterious variants in one gene co-occur with deleterious variants in a specific second MES gene (always in concert with common genetic variation and environmental factors), their cumulative contributions can cross the liability threshold. This does not imply that gene pairs are epistatic or form dependent gene-gene interactions. Instead, we find paired genes are predominantly independent with co-occurrence being strongly associated with ASD. Thus, our strategy targets genes with meaningful but not singular impacts on ASD liability.

**Figure 1.**
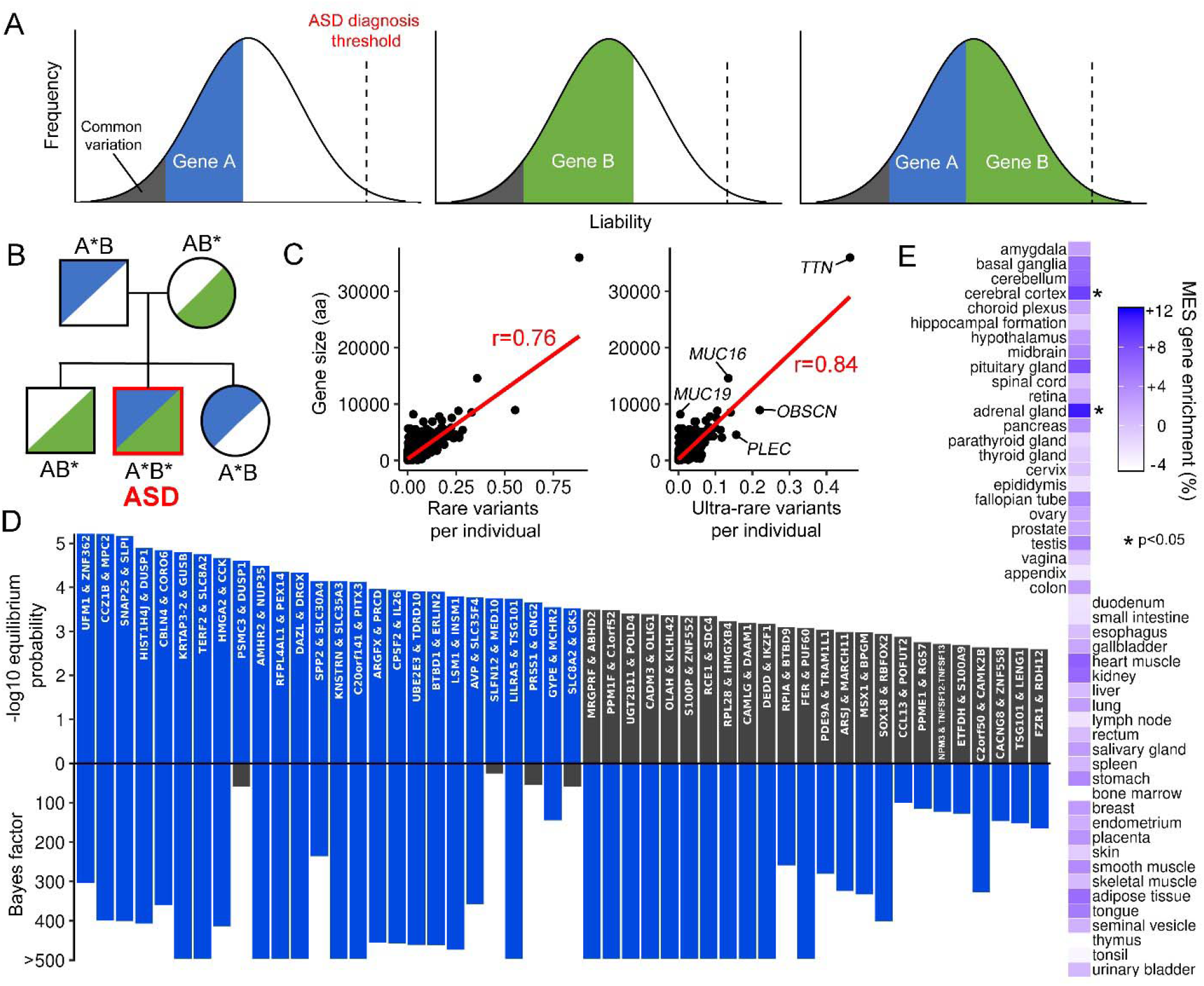
Discovery and enrichment of MES genes. (A) Hypothesized role of individual MES genes and their cumulative influence on the genetic liability of ASD in an individual. (B) Example inheritance pattern illustrating ultra-rare deleterious variants in two genes within the parents (depicted in blue or green) that are exclusively jointly inherited by offspring with ASD. This exemplifies a potential genetic mechanism contributing to ASD susceptibility. (C) Mean number of rare (<1%) or ultra-rare (<0.1%) deleterious variants in SPARK parents versus gene size. The red line represents linear regression. Without *TTN*, correlation is 0.71 and 0.80, respectively. (D) Equilibrium probability and Bayes factors for the predicted MES gene pairs in ASD. Bars in blue denote an equilibrium probability <3.03x10^-4^ or a Bayes factor >100. (E) Enrichment of predicted MES genes per tissue relative to all protein-coding genes.

The process of testing gene combinations poses a statistical challenge, particularly given the vast number of potential gene pairs. For example, with 20,000 protein-coding genes, the number of two-gene combinations is approximately 200 million, necessitating an extremely stringent significance threshold. To overcome this challenge, we selected candidate pairs based on familial inheritance patterns of ultra-rare deleterious variants, narrowing the focus to a manageable subset of potential risk genes. Subsequent statistical analysis on this subset aimed to detect gene pairs more prevalent in individuals with ASD compared to non-ASD controls. After establishing statistical significance and validating findings, we then characterized 97 predicted MES genes to further support their roles in ASD etiology and heterogeneity.

## Results

### Identification of candidate MES genes inherited as pairs

Our search for MES genes began by identifying pairs of candidate genes through family-based inheritance patterns. This approach circumvents conducting approximately 200 million pairwise comparisons among all protein-coding genes which would impose a harsh significance threshold set by multiple testing corrections. Our primary objective was to identify instances where inherited variants in two distinct genes were overrepresented in offspring with ASD, with non-ASD offspring potentially inheriting either variant, but not both. This streamlined approach enabled us to focus on candidate gene pairs exhibiting the most promising associations with ASD.

The first and second steps of our candidate screen focused on the identification of familial structures wherein ultra-rare (frequency <0.1%) deleterious variants in two genes were inherited by all offspring with ASD within a given family (**Fig 2**; **Fig 1B**). In this context, the term ‘A*’ designates an ultra-rare deleterious genetic alteration in a gene present in one parent, with the potential for transmission to the offspring. The second step searched for mirrored inheritance patterns involving an ultra-rare deleterious variant in a different gene, denoted as ‘B*’. In such instances, ‘B*’ originates from the parent who did not transmit ‘A*’, thereby maintaining genetic independence. We then filtered to instances where all ASD offspring that share the same parents exhibited ‘A*B*’ inheritance, while none of their non-ASD siblings carried both alleles. In different families, the specific ultra-rare deleterious variants in genes were predominantly distinct. Additionally, as there were instances of multiple ‘A*’ or ‘B*’ variants within the same gene and family, we used the count of unique families rather than unique sites for identifying ‘A*B*’ pairs. We initially analyzed 10,795 families from the SPARK^15^ iWES v1 dataset (“SPARK v1”, comprising waves WES1-WES4), each with both parents and at least one ASD offspring genotyped with whole exome sequencing data (WES; see **Methods**). For deleterious ultra-rare variants (gnomAD^16^ v3 non-neuro allele frequency of <0.1%), families on average exhibited an ‘A*’ inheritance pattern for 94 different genes.

**Figure 2.**
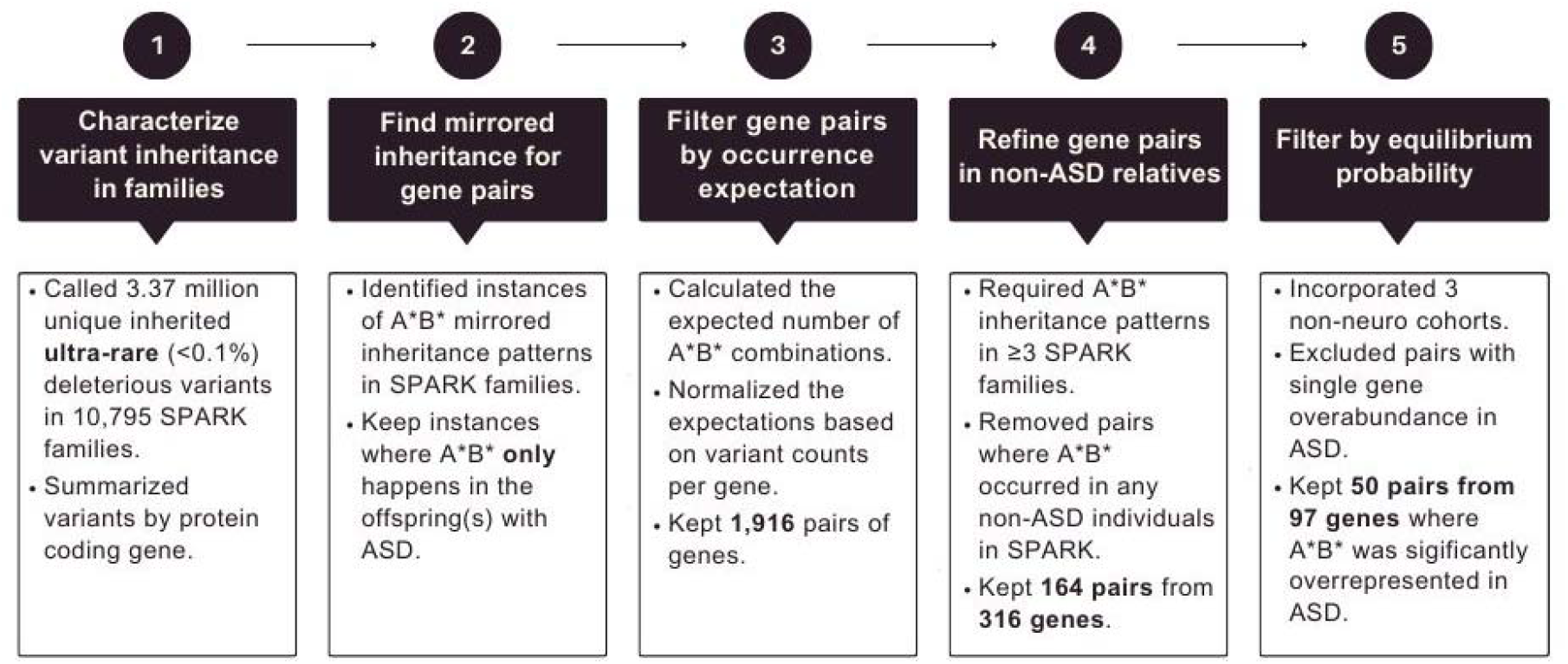
Flowchart of steps involved in MES gene discovery.

In the third step, to identify promising gene pairs for further analysis, we assessed the likelihood of observing specific patterns of mirrored ‘B*’ inheritance within families displaying ‘A*’ inheritance. This entailed calculating the likelihood of ultra-rare deleterious ‘B*’ variants manifesting in a second gene, contingent upon our knowledge of the prevalence of such variants in the population. For instance, let us consider ‘GeneA’ which features ‘A*’ inheritance in seven families, and ‘GeneB,’ another gene with ‘B*’ inheritance in 20 families. Among these, four families exhibited the aforementioned mirrored ‘A*B*’ inheritance pattern. Utilizing the binomial distribution and adjusting by the frequency of ultra-rare deleterious variants per gene in the SPARK parent population, we determined (1) the expected frequency of observing this four-in-seven and, conversely, four-in-twenty arrangement, and (2) the relative likelihood that the genes observed were ‘GeneA’ and ‘GeneB’ (see **Methods**). This latter step is essential as genes with longer coding sequences generally have a greater frequency of ultra-rare or rarer deleterious variants (Pearson’s *r*: 0.84; 0.76 for rare or rarer variants; **Fig 1C**). To illustrate, ‘A*’ inheritance patterns within the large gene *TTN* were observed in 32.0% of all SPARK families. Consequently, the occurrence of a mirrored ‘A*B*’ inheritance pattern with *TTN* is considerably more anticipated than with *SNAP25*, a small gene where ‘A*’ inheritance patterns were present in only 0.048% of the SPARK families.

We retained 1,916 pairs of genes where the normalized expected frequency was less than one in both ‘A*’ and ‘B*’ configurations (**Table S1**). Additionally, in the fourth step, for each candidate pair, we imposed the condition that at least three families must exhibit the ‘A*B*’ inheritance pattern. To refine candidate pairs, we required that there were no occurrences of ‘A*B*’ in the remaining non-ASD individuals within the broader SPARK dataset, which comprised 36,323 individuals sequenced with WES. This filtering process resulted in a final count of 164 candidate pairs from 316 unique protein coding genes (**Table S1**).

Of note, we tested additional variant types and frequencies, including missense variants and PTVs of rare (<1%) frequency and analyses restricted to only PTVs (see **Methods**). We selected PTVs and missense variants of ultra-rare or rarer frequency for the following analyses as it produced the greatest number of candidate pairs after SPARK family filtering.

### Statistical assessment of MES genes and incorporating non-neuro cohorts

Though SPARK includes individuals without ASD, all are within families where at least one individual has ASD. This makes these individuals an inaccurate representation of the general population. Therefore, to perform statistical analyses below that incorporate the population frequency of ultra-rare deleterious variants by gene, we included individuals without ASD or other known mental, behavioral, or neurodevelopmental disorders. This included 3,202 individuals from the 1000 genomes project (1kGP)^17,18^, 156,550 individuals from the All of Us (v7) consortium, and 16,586 individuals from the BioMe Biobank. As with SPARK, we identified ultra-rare deleterious variants in the 316 candidate MES genes in these cohorts. Lacking familial information for most individuals, we could not determine with certainty whether any variant was inherited or *de novo*. However, given that there are approximately 70 *de novo* mutations per genome per generation^19^, we can assume that variants observed in these cohorts are almost always inherited.

To ensure comparability of detected variants in the 1kGP, All of Us, and BioMe cohorts as controls, we examined the frequency of ultra-rare deleterious variants within the candidate MES genes across these cohorts. Positive correlations were observed among all cohorts, with a more pronounced correlation in cohorts characterized by larger sample sizes (**Fig S1**). Notably, non-ASD individuals in the SPARK cohort displayed a slightly higher burden of ultra-rare deleterious variants per gene compared to individuals in the other cohorts. On average, candidate genes in the SPARK cohort contained 0.02%, 0.12%, and 0.18% more variants than in the All of Us, BioMe, and 1kGP cohorts, respectively. Across all cohorts, SPARK showed a notable and consistent enrichment for ultra-rare deleterious variants in *UGT2B11*, *AFAP1L2*, *C20orf141,* and *OLAH* (**Fig S1**). However, these genes also contained relatively more synonymous variants in the SPARK cohort, suggesting their enrichment for deleterious variants is not biologically meaningful. Of note, variants in Z*NF717* were highly enriched in 1kGP compared to the other cohorts (5.74-8.78% more). However, this gene’s role in cellular proliferation and the lymphoblastoid cell line source of 1kGP suggest these variants may be somatic and under positive selection.

Among the 212,661 individuals without ASD, ‘A*B*’ was conspicuously absent in 26 out of the 164 candidate pairs and occurred in only two or fewer individuals in 83 candidate pairs. Across all candidate pairs, the incidence of ‘A*B*’ was consistently higher among individuals with ASD than their non-ASD counterparts. Notably, in 71 pairs, the frequency of ‘A*B*’ was more than 10 times greater in individuals with ASD. Together, this illustrates a pronounced disequilibrium between individuals with ASD and those without ASD.

Our above search (encompassing steps one through four in **Fig 2**) aimed to identify potential MES genes for in-depth analysis, mitigating the influence of large genes with a high frequency of ultra-rare deleterious variants. In the fifth and final step, our objective was to assess the statistical significance of the skew in MES gene pairs in relation to ASD. We hypothesized that the concurrent inheritance of ultra-rare deleterious variants in two paired MES genes was strongly associated with ASD development. We anticipated that the prevalence of ‘A*B*’ in the population would adhere to an expected equilibrium, being the product of ‘A*’ and ‘B*’ frequencies in the population. Their significant enrichment in individuals with ASD is presumably the result of their involvement in ASD etiology. In contrast, our null hypothesis is that the overrepresentation of ‘A*B*’ in individuals with ASD, contingent upon the individual frequencies of ‘A*’ and ‘B*’ in the population, occurred by chance. Furthermore, when examining the frequency of ‘A*’ or ‘B*’ in isolation, we required that neither gene may be significantly overrepresented (p<0.05) in the individuals with ASD. Therefore, with respect to ASD, we required only ‘A*B*’ to be in strong disequilibrium.

To illustrate, consider ultra-rare deleterious variants in the genes *SNAP25* (as ‘A*’) and *SLPI* (as ‘B*’), which are present in 0.056% and 0.196% of individuals, respectively. Notably, these frequencies do not show significant differences when queried in individuals with or without ASD (0.053% versus 0.057% for *SNAP25* and 0.220% versus 0.192% for *SLPI,* respectively). At equilibrium, ‘A*B*’ should occur at a frequency determined by the product of their individual frequencies. For *SNAP25* and *SLPI,* ‘A*B*’ occurs in three of 34,164 individuals with ASD and in none of the 212,661 individuals without ASD. Using the cumulative density function of the binomial distribution, we calculated the probability of observing at least three individuals with ASD having ‘A*B*’ and none without ASD to be 6.88x10^-6^. We calculated this equilibrium probability for all 164 paired candidate MES genes. For 33 pairs, ‘A*B*’ was significantly overrepresented in individuals with ASD (p<3.03x10^-4^, corrected for multiple testing). We discarded seven candidate pairs where an individual gene in isolation was significantly overrepresented in individuals with ASD.

We additionally calculated the Bayes factor for each pair. As described above, our null hypothesis posits that the increased occurrence of ‘A*B*’ in individuals with ASD is a random event occurring at the equilibrium probability observed above. Our alternate hypothesis is that ‘A*B*’ is enriched among individuals with ASD precisely because it results in an ASD phenotype. Therefore ‘A*B*’ would be more at equilibrium when not considering ASD status. Using the example of *SNAP25* and *SLPI*, the Bayes factor was 400, strongly supporting the alternate hypothesis. Interestingly, one of the largest Bayes factors was 5,807 for the genes *PITX3,* a gene with a known role in dopaminergic neuron differentiation, and *C20orf141*, a gene of unknown function. Across the 164 paired candidate MES genes, 46 had a Bayes factor greater than 100.

We retained 50 candidate gene pairs (97 unique genes) where the equilibrium probability was <3.03x10^-4^ or the Bayes factor was >100 (**Fig 1D**; **Table S1**). Importantly, the genes in pairs showed statistically significant associations to ASD but not when tested in isolation. From here, we will refer to these as ‘predicted MES genes’.

According to the recent and most comprehensive screens for ASD genes^4^, the mean ASD FDR of the predicted MES genes was 0.804, demonstrating that single MES genes by themselves are not generally associated with ASD. Only one gene, *DEDD*, had an ASD FDR <0.05. Six genes (*PUF60, SNAP25, PSMC3, CAMK2B,* and *HIST1H4J*) were associated with neurodevelopmental disorders (FDR <0.05) though not for ASD. Similarly, the 97 MES genes were also not associated with ASD in other major studies ^3,10,11,20^ and are therefore not considered consensus ASD-associated genes.

The median probability of being loss-of-function intolerant (pLI) score of the predicted MES genes was 0.029 and only 15 genes were >0.9, the common threshold for extreme haploinsufficiency. When breaking up genes by probability of pLI decile, MES genes inhabited more moderate deciles than large effect size genes characterized in the most recent comprehensive assessment of ASD genes ^4^ (**Fig S2**). This was also the same result for loss-of-function observed over expected upper bound fraction (LOEUF) scores (**Fig S2**). This suggests the predicted MES genes are not strongly constrained and are more haplosufficient. This is again consistent with our model of MES genes where each gene alone is not evidently associated with ASD. Importantly, deleterious variants in both copies of predicted MES genes are not present nor assessed in this study. Loss-of-function in both copies of a MES gene could produce a separate pathological phenotype. Nonetheless, we present 97 predicted MES genes that when disrupted in concert with a second MES gene are strongly associated with ASD.

### Validations of MES genes

To mitigate the challenge of a highly stringent multiple testing correction threshold inherent in comparing every protein-coding gene, we streamlined our analysis to focus on candidate gene pairs. This approach, while efficient, raises concerns of bias due to selective inference, as it prioritizes pairs of genes with a pre-established likelihood of association. To address potential bias, we implemented two validation strategies: (1) conducting a replication study using synonymous variants as a neutral benchmark, and (2) incorporating additional ASD samples.

Our initial validation utilized ultra-rare synonymous variants, which, due to their non-functional impact, were anticipated to yield fewer or less significant associations with ASD. This analysis was conducted identically to that for deleterious variants. We found a similarly strong correlation between the quantity of ultra-rare synonymous variants and CDS length (Pearson r: 0.83), as well as between the counts of ultra-rare synonymous and deleterious variants (r: 0.82; **Fig S3 A, B**). Moreover, the variability in the frequency of ultra-rare synonymous variants across different cohorts paralleled that observed for missense variants (**Fig S3C**), affirming that synonymous variants replicate our analyses under the same conditions. Utilizing ultra-rare synonymous variants, we identified merely five gene pairs (10 unique genes; **Table S1**), in contrast to the 50 pairs identified with deleterious variants. Together, this suggests a potential false-positive rate of approximately 10%.

Further validation involved a new cohort of 36,997 samples from SPARK iWES v2 (“SPARK v2”, adding WES5), including 10,128 ASD individuals. Applying the same methodological framework, we first reassessed the equilibrium tests using only the new SPARK samples. However, only eight of the original 50 gene pairs could be validated as we required A*B* in at least one individual with ASD. Among the eight, seven pairs met the criteria of an equilibrium probability p<0.05 or a Bayes Factor >10, adjusted for the smaller independent dataset (**Fig S4A**). None of the five pairs identified with ultra-rare synonymous variants could be validated. In addition, by combining data from both the SPARK v1 and v2 cohorts, we found 38 of the original 50 pairs maintained significance (equilibrium probability <3.03x10^-4^ or a Bayes factor >100; **Table S1; Fig S4B**). By contrast, only two synonymous variant pairs retained significance post-combination, none surpassing a significance level of 1.2x10^-4^, a stark contrast to the maximum significance of 5.9x10^-8^ observed in the primary analysis with ultra-rare deleterious variants (**Table S1**).

To summarize, our methodology efficiently identifies MES genes while maintaining a low false-positive rate. Of the initial 50 gene pairs, 38 (74 unique genes) were corroborated by integrating the SPARK v2 dataset, with seven out of eight subjected to independent validation using SPARK v2. Subsequent analyses will delve into the 97 unique genes within these pairs, focusing on their expression patterns and phenotypes among carriers as further evidence of their involvement in ASD.

### Enrichment of MES gene expression in the brain

To further our identification of MES genes implicated in ASD, we conducted an analysis of their tissue expression profiles. While ASD etiology encompasses factors beyond neural influences, such as those associated with the gut microbiome ^21^, it is imperative that genes contributing to ASD development exhibit expression in the brain. Accordingly, we hypothesized that our predicted MES genes would exhibit an enrichment for expression in brain tissues. Utilizing consensus normalized expression values (nTPM) from the Human Protein Atlas (v23)^22^, and encompassing 50 tissues, we observed that 77.0% of the 20,151 protein coding genes demonstrated detectable expression (nTPM > 1) in at least one brain tissue, with 70.5% exhibiting expression in the cerebral cortex. Strikingly, among the 97 predicted MES genes, 85.6% displayed detectable expression in at least one brain tissue, and 79.4% specifically in the cerebral cortex. To assess the statistical significance of these findings, we performed random gene sampling and quantified per-tissue expression, revealing a significant overrepresentation of predicted MES genes in the cerebral cortex (p=0.022) and the adrenal glands (p=0.009) (**Fig 1E**; **Fig S5A**). The predicted MES genes did not exhibit significant underrepresentation in any tissues. Among the 74 unique genes that maintained significance with the addition of SPARK v2, we observed the same cerebral cortex and adrenal gland enrichment. Furthermore, we did not observe that the predicted MES genes were expressed at a significantly higher level than other cerebral cortex or adrenal expressed genes. Nonetheless, the observed enrichment of MES gene expression in the cerebral cortex aligns with our initial predictions. The enrichment in the adrenal glands was unexpected but not unprecedented as increases in androgen and cortisol levels, which are both secreted by the adrenal glands, are associated with ASD ^23,24^.

To conduct a more in-depth investigation, we queried gene expression within 193 subregions of the brain (**Fig S5B**). Notably, we identified the most pronounced enrichment of the 97 predicted MES genes in the ventral tegmental area and central amygdala, although the statistical significance fell just below the conventional threshold (p=0.08 and p=0.07, respectively). Again, the same pattern was observed with the 74 unique genes that maintained significance with SPARK v2. It is noteworthy that these particular brain regions exhibited the highest proportion of MES genes with detectable expression (83.5% for both regions), surpassing the expression levels observed in other brain regions or any alternative tissues that we analyzed. The 97 predicted MES genes are not canonically associated with ASD because they do not show strong associations when tested alone. Yet, their expression is most pronounced in a region of the brain linked to psychiatric disorders including ASD ^25,26^. Together, we showed the predicted MES genes are characterized by expression in neural tissues, suggesting their disruptions may result in psychiatric phenotypes.

### Phenotypes of non-ASD carriers of MES genes

While MES genes were identified in pairs, the nature of their contribution to ASD remained ambiguous – whether each gene within a pair independently influenced ASD or, conversely, if their impact stemmed from interdependent gene-gene (GxG) interactions. In the scenario of independent contributions, the disruption of a single MES gene may manifest mild ASD-like phenotypes, while the disruption of two MES genes could result in an additive or synergistic effect. In contrast, within the context of dependent GxG interactions, the disruption of a single MES gene yields no discernible phenotype. To observe if any predicted MES pairs show independent contributions, we queried the phenotypes and questionnaire responses of non-ASD individuals within the SPARK v1 cohort. Per our MES gene discovery methodology, non-ASD individuals may have an ultra-rare deleterious variant in either gene (herein simply referred to as ‘carriers’) of an MES gene pair – but never both.

We first examined responses from the Social Communication Questionnaire (SCQ), completed for adolescent offspring with and without ASD. The SCQ, comprising 40 yes-or-no questions designed for ASD screening, was available for 3,831 non-ASD offspring in our study. We focused on 68 out of the 97 predicted MES genes that exhibited a minimum of five non-ASD MES gene carriers with SCQ responses. For each gene and each SCQ question, we employed a χ test to compare responses between carriers and non-carriers among the non-ASD offspring. We found 31 out of the 68 tested genes exhibited a significant (p<0.05) skew for at least one SCQ question. For 15 genes, more than one question exhibited significantly increased ASD-like responses, with a maximum of six questions per gene. In total, 65 questions demonstrated significant skew, with 63 revealing a higher frequency of ASD-like responses in carriers compared to non-carriers (**Fig S6**). Our results remained consistent when comparing carriers to the 3,007 non-ASD offspring devoid of ultra-rare deleterious variants in any of the 97 predicted MES genes (65 of 70 significant questions indicated carriers with heightened ASD-like responses).

In addition to examining individual SCQ questions, we assessed the total SCQ score. No discernible elevation in total scores was observed among carriers of any MES genes. The most substantial mean score difference between non-ASD carriers and non-carriers was less than two points. Nonetheless, our study demonstrated that non-ASD carriers of MES genes may exhibit significant differences in responses to specific SCQ questions, but not in overall scores. Nevertheless, our results suggest that individual MES genes can contribute to a mild social deficiency phenotype.

Though ASD-related phenotype measurements were less abundant for non-ASD individuals, we were able to evaluate the presence of other psychiatric or developmental conditions through the basic medical questionnaire. Across 28,883 non-ASD individuals and 29 questions on the presence of developmental or psychiatric conditions (e.g., schizophrenia, obsessive-compulsive disorder, sleep disorders, motor delays), we found 47 of 97 tested MES genes exhibited a significant (p<0.05) skew for at least one condition (**Fig S7**). In 83.6% of cases, carriers of the MES gene showed increased incidence of the condition. For example, 15.5% of non-ASD carriers of ultra-rare deleterious variants in *PITX3* reported obsessive-compulsive disorder (OCD) compared to 4.3% of non-carriers (p=0.004). Of note, the minority of cases where MES gene carriers had significantly lower incidence occurred exclusively for conditions that were relatively common (>5% of non-ASD respondents) such as depression and social anxiety. Again, these results were consistent when comparing to non-ASD offspring devoid of ultra-rare deleterious variants in any of the 97 predicted MES genes. In summary, we found evidence that many individual MES genes may contribute independently to specific ASD-related traits and conditions in non-ASD carriers.

### Phenotypes of ASD carriers of MES genes

The heightened prevalence of psychiatric conditions in non-ASD carriers of individual MES genes suggests a potential influence on the diverse presentation of ASD. It is conceivable that MES genes contribute to both the overall liability of ASD and the liability of comorbid symptoms associated with ASD. In the SPARK v1 cohort, 99.95% of individuals with ASD exhibit at least one of the 29 comorbid disorders assessed. Notably, language delays are the most common, affecting 51% of individuals, followed by attention-deficit/hyperactivity disorder (ADHD) at 38%.

Among 58 of the 97 MES genes analyzed, carriers with ASD displayed significantly altered frequencies of comorbid disorders compared to ASD individuals without any MES genes (**Fig 3A**). For 31 MES genes, comorbid disorders were always more prevalent. For instance, individuals with ASD carrying an ultra-rare deleterious variant in the gene *NPM3* demonstrated a significantly higher incidence of motor delay (p=0.002), diagnosed sleep disorders (p=0.007), separation anxiety (p=0.02), social communication disorder (p= 0.02), eating disorders (p=0.02), and encopresis (p=0.03). Importantly, carriers of *NPM3* among non-ASD individuals also exhibited a significantly higher incidence of learning disabilities (p=0.01) and separation anxiety (p=0.04). In total, among the 31 MES genes associated with increased comorbidity frequency in individuals with ASD, 14 were also linked to increased comorbidity frequency in non-ASD individuals, though not always for the same comorbidity.

**Figure 3.**
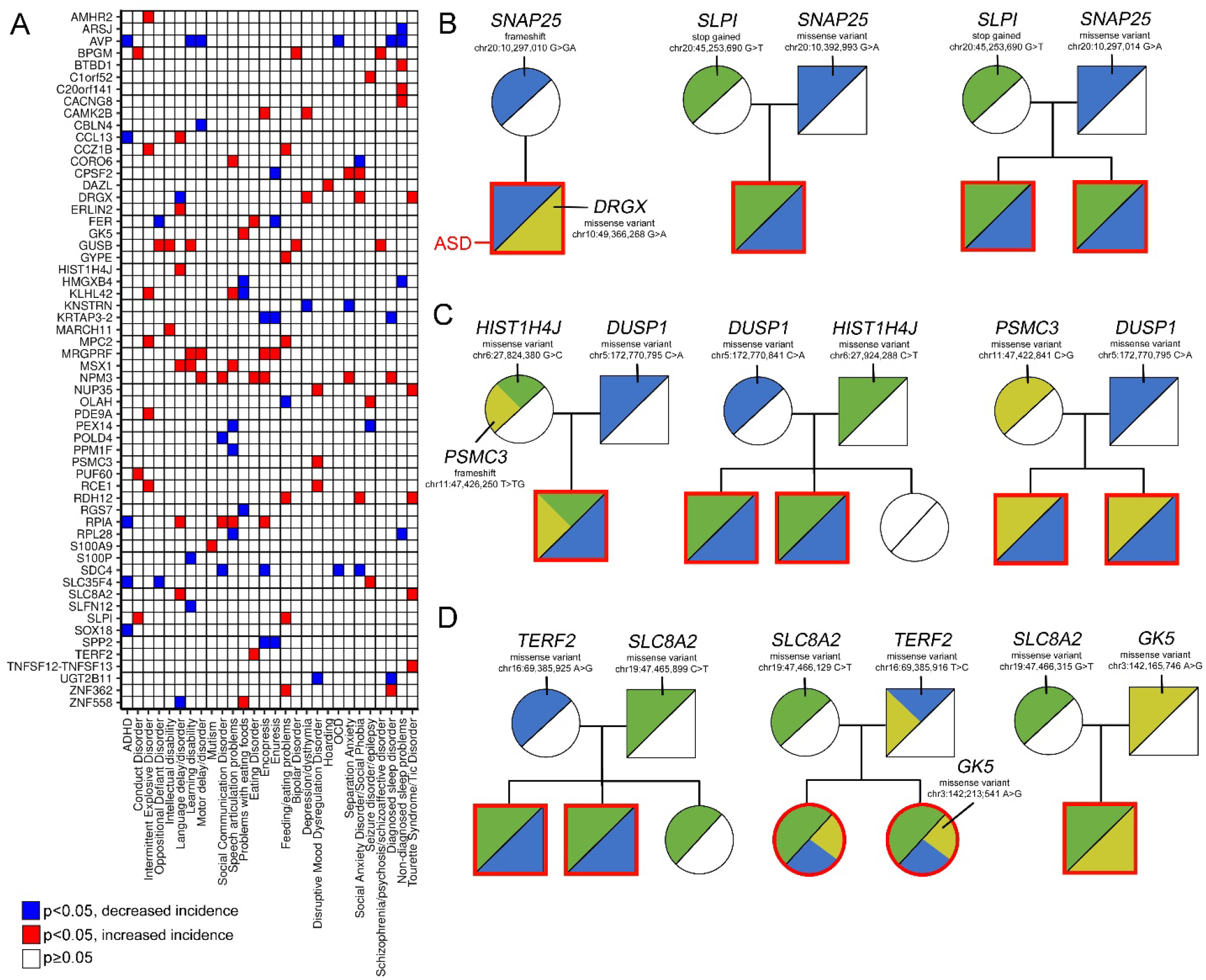
Phenotypes and familial inheritance of MES genes. (A) Differences in comorbidity frequency of carriers of MES genes with ASD. Significant differences in ASD carrier versus non-carrier condition frequencies are highlighted. Only genes with at least one significant comorbidity bias are shown. (B) Example families from SPARK v1 that carry mutations in *SNAP25.* (C) As in panel B for *DUSP1*. (E) As in panel B for *SLC8A2*.

Interestingly, for 17 MES genes, individuals with ASD exclusively exhibited significantly lower incidence of comorbidities (**Fig 3A**). Carriers of ultra-rare deleterious variants in *AVP*, the gene responsible for the production of the neuropeptide hormone arginine vasopressin, demonstrated a reduced incidence of ADHD (p=5x10^-4^), sleep disorders (p=0.004), OCD (p=0.03), motor delays (p=0.03), and learning disabilities (p=0.048). Notably, in non-ASD individuals, carriers of *AVP* exhibited a decreased incidence of depression (p=0.02) and a noteworthy, albeit not statistically significant, decrease in ADHD and sleep disorders. As *AVP* is anxiogenic and promotes the stress response ^27^, we tangentially hypothesize its disruption could reduce these effects. In addition, a further 10 MES genes showed increased frequencies for one comorbidity while decreased frequency for another. Carriers of *DRGX*, a gene involved in nervous system development, showed elevated incidence of social anxiety disorder (p=0.03), depression (p=0.049), and Tourette Syndrome (p=0.04) but a decreased incidence of language delay (p=0.01). Thus, we find evidence MES genes can both contribute to the genetic liability of ASD and its heterogeneity.

### Co-expression and interactions of MES gene pairs

Above, we demonstrated that 76 of the 97 identified MES genes exhibited discernible phenotypic associations, suggesting most MES genes do not form dependent interactions when contributing to ASD risk. To further test dependence, we inquired into the co-expression and protein interaction patterns of these MES gene pairs compared to random MES gene pairings.

Employing the tissue-based expression data used above, we found that for 48 of 50 gene pairs, the correlation in tissue co-expression did not significantly surpass that observed between any two MES genes (**Fig S8A**). Notably, only the paired genes *CADM3* and *OLIG1* demonstrated a significantly elevated co-expression correlation (p=0.028; Pearson’s r= 0.46). However, carriers of *CADM3* demonstrated altered SCQ responses, suggesting the pair’s high co-expression is not compelling evidence of dependence. Interestingly, the other gene pair with significant co-expression, *CACNG8* and *ZNF558,* was anti-correlated (p=0.032; r= -0.38). Using the expression profiles of 193 brain subregions, we also found three pairs demonstrated significantly elevated co-expression and three pairs with significant anti-correlation (**Fig S9A**).

Interestingly, one of these significantly anti-correlated pairs was *CADM3* and *OLIG1* (p= 0.012, r= -0.72) indicating their cross-tissue expression pattern was alike while their brain subregion pattern was opposing. Together, we do not find evidence that paired MES genes are more co-expressed than any two MES genes, suggesting the pairs predominantly contribute independently to ASD risk.

We additionally clustered MES co-expression patterns and compared them to 185 high confidence ASD genes^4^. We found that MES genes did not cluster separately from these established ASD genes, suggesting that MES genes integrate into similar tissue-expression pathways (**Fig S8B**). This same finding was also observed using the expression from 193 brain subregions (**Fig S9B**).

We additionally investigated protein-protein interactions between paired MES genes using STRING (v12)^28^. Of the 97 MES genes, 40 interacted with some second MES gene with medium confidence (interaction score ≥0.4). However, only one MES pair, *HMGA2* and *CCK*, showed evidence of interaction (score=0.44). This again supports our hypothesis that MES pair genes do not form dependent interactions.

### Illustrative examples of MES gene pairs

Within this investigation, we have delineated the identification of 97 MES genes in 50 pairs, exploring their connections to brain expression patterns and ASD-like phenotypes. To conclude, we will highlight specific MES genes in these pairs, with particular attention given to familial case studies. These cases not only underscore the significance of inheriting multiple MES genes but also highlight the disparate contributions of MES genes to the genetic susceptibility of ASD.

The concurrent inheritance of ultra-rare deleterious variants in *SNAP25* (Synaptosome Associated Protein 25) and *SLPI* (Secretory Leukocyte Peptidase Inhibitor) emerged as a robust association with ASD (equilibrium probability of 6.9x10^-6^; Bayes factor of 400). *SNAP25,* a synaptic protein, is one of the highest expressed genes in the brain. While variants in *SNAP25* have been linked to ADHD, schizophrenia, bipolar disorder, and neurodevelopmental disorders, it often fails to be linked to ASD ^3,4,29,30^. Our findings support this inconsistency as loss-of-function disruptions of one copy of *SNAP25* did not always coincide with ASD, best exemplified by an inherited frameshift variant in a non-ASD mother transmitted to her child with ASD (**Fig 3B**). This suggests that meeting the genetic liability for ASD among *SNAP25* carriers necessitates additional contributions from common variations or another MES gene. In the case of the mother-to-son transmitted frameshift in *SNAP25*, the ASD-afflicted child, but not the mother, harbored an ultra-rare deleterious variant in another MES gene, *DRGX*, implicated in nervous system development. *SLPI*, another candidate paired with *SNAP25* (**Fig 3B**), is integral to the innate immune system and serves a protective role against inflammation in tissues ^31^. Notably, this candidate assumes relevance given the well-established association between inflammatory conditions and ASD ^32,33^.

Another noteworthy candidate MES gene is *DUSP1* (Dual Specificity Phosphatase 1), a gene implicated in the regulation of inflammation and nervous system development ^34^. Our investigation revealed a strong association of *DUSP1* with ASD when co-occurring with *HIST1H4J* (H4 Clustered Histone 11; *H4C11*) or *PSMC3* (Proteasome 26S Subunit, ATPase 3), with both genes linked to intellectual disability and neurodevelopmental delay ^35,36^. Interestingly, the concurrent inheritance of ultra-rare deleterious variants in *PSMC3* and *HISTH4J* was observed in only two individuals—a non-ASD mother and her son with ASD (**Fig 3C**). Though a limited occurrence, this raises the possibility that *PSMC3* and *HISTH4J* may have comparatively lesser contributions to the genetic liability of ASD when compared to *DUSP1*. This inference is reinforced by the familial context, as the son with ASD inherited a *DUSP1* variant from the father in addition to the *PSMC3* and *HISTH4J* variants from the mother. This unique familial pattern suggests a more prominent role for *DUSP1* in the manifestation of ASD within this specific genetic context.

The last candidate MES genes we will illustrate are *SLC8A2* (Solute Carrier Family 8 Member A2) which associates with ASD when paired with either *TERF2* (Telomeric Repeat Binding Factor 2) or *GK5* (Glycerol Kinase 5) (**Fig 3D**). *SLC8A2*, a sodium-calcium exchanger, is highly expressed in neurons where it plays a role in sodium and calcium ion homeostasis ^37^. *TERF2* plays an important role in telomere maintenance and neuronal differentiation ^38^ whereas *GK5* is a broadly expressed gene that is part of glycerol metabolism. The concurrent inheritance of ultra-rare deleterious variants in *TERF2* and *GK5* was observed in four individuals – two with and two without ASD. Three of these individuals were in the same family where variants in both genes were transmitted from a non-ASD father to his two daughters with ASD (**Fig 3D**).

However, as was observed above with *DUSP1, PSMC3,* and *HISTH4J,* the daughters with ASD also inherited variants in *SLC8A2* from the mother. This case study again suggests that *SLC8A2* has a greater contribution to the genetic liability of ASD than *TERF2* and *GK5.* In summation, the interplay of our predicted MES genes underscores the polygenic landscape of genetic factors contributing to ASD susceptibility and heterogeneity.

## Discussion

This study presents a novel approach to understanding the complex genetic landscape of ASD by identifying MES genes – genes that individually, when harboring a deleterious variant, fall below the ASD liability threshold but, when paired with another MES gene, collectively contribute to ASD risk. By analyzing familial inheritance patterns of ultra-rare deleterious variants, we identified candidate pairs of genes further assessed in 255,883 individuals of which 31,183 have ASD. We revealed 97 predicted MES genes forming 50 pairs, with significant associations to ASD when considered together but not individually. This study emphasized the importance of gene combinations, shedding light on the nuanced interplay of genetic factors in ASD susceptibility and providing insights into the disorder’s heterogeneity. Additionally, we explored the expression profiles of MES genes in the brain and examined the phenotypic effects of carriers, offering a comprehensive perspective on the multifaceted genetic contributions to ASD.

Our study focused on identifying a novel but limited set of paired MES genes with the most pronounced associations to ASD. A primary limitation of this study is that we did not consider common genetic variation or environmental conditions, assuming that the co-inheritance of paired MES genes alone was sufficient to result in ASD. However, it is well-established that common genetic variation and environmental factors substantially contribute to ASD liability. Further analyses of MES or even large effect size genes may involve comparing carriers with high or low polygenic risk scores, potentially revealing that single MES genes and a high burden of common variation may be adequate to cross the diagnostic threshold. The incorporation of common variation may also aid in uncovering even smaller effect size genes. Detecting combinations of three or more genes using our methodology would necessitate a larger number of families. However, identifying smaller effect size genes conditioned on a high or low polygenic risk score remains an avenue for future exploration.

Our investigation sought evidence for whether identified MES genes act independently or participate in dependent GxG interactions to influence ASD susceptibility. SCQ responses among non-ASD carriers revealed a significant skew for specific questions, suggesting that many individual MES genes may independently contribute to subtle social deficiencies. Furthermore, altered frequencies of developmental or psychiatric conditions in both non-ASD and ASD carriers supported the notion that MES genes can independently influence diverse phenotypic outcomes. In total, 76 out of the 97 predicted MES genes exhibited skewed frequencies in at least one SCQ response or the incidence of a developmental or psychiatric condition. It is plausible that some MES gene pairs form dependent GxG interactions, implying carriers of one gene exhibit no discernable differences from non-carriers. Notably, there were instances where non-ASD individuals harbored ultra-rare deleterious variants in up to five MES genes. MES gene burden in non-ASD individuals was not associated with sex at birth, suggesting this is not a result of the female protective effect. Instead, in cases where non-ASD individuals possessed more than two MES genes, the 21 genes without a skewed frequency in SCQ or developmental/psychiatric conditions constituted an average of 90% of the multiple MES genes per individual. If dependent GxG interactions describe this minority of genes, it would allow non-ASD individuals to carry multiple unpaired MES genes without affecting their phenotype. This nuanced understanding of the independent and collective actions of MES genes provides insights into the intricate nature of their contributions to ASD susceptibility, unraveling the complex interplay between genetics and phenotypic expression.

We hypothesized that MES genes would exhibit less conservation than larger effect size genes which was indeed reflected by their moderate pLI deciles. In the context of ASD, genes with large effect sizes typically manifest autosomal-dominant effects, leading to profound phenotypes such as epilepsy, developmental delay, and intellectual disability, alongside ASD ^3^. Consequently, *de novo* mutations in these genes are seldom transmitted to subsequent generations. In contrast, loss-of-function mutations in MES genes tend to result in less severe phenotypes if any, possibly due to redundant pathways or functional paralogs. As a result, disruptions in these genes can be transmitted and still contribute to the heritability of ASD. The high heritability of ASD implies that its key genetic factors may generally be subject to more lenient evolutionary constraints. Variants in genes that tolerate altered function and expression may follow similar trends. The ability of MES genes to tolerate mutations and still contribute to ASD’s heritability underscores the importance of understanding the interplay between genetic variation, phenotypic outcomes, and evolutionary pressures in the context of ASD susceptibility.

The most challenging aspect of MES genes is that the disruption of a single gene is generally inadequate to induce ASD, yet still contributes to its etiology and heterogeneity. This necessitates a nuanced approach in determining how terms such as “causal genes,” “risk genes,” or “pathogenic variants” are to be applied. In the context of monogenic diseases, the causality of a gene or mutation is often straightforward and direct. However, the causality associated with complex diseases like ASD entails a combination of multiple genetic variants and environmental factors contributing to the progression of the condition, rather than merely elevating the risk. Although a MES gene may be considered causal as it influences key pathways in ASD development, it does not singularly determine the condition. Guidelines, such as those established by the American College of Medical Genetics and Genomics (ACMG), play a pivotal role in interpreting and categorizing genetic variants, ranging from pathogenic to benign. ACMG incorporates an array of factors, including functional predictions, inheritance patterns, and prior reports, to assign pathogenicity to genes or variants ^39^. This approach is highly effective at identifying strong gene drivers of human diseases but less applicable to complex conditions where MES genes may play crucial roles. Variants in MES genes may be assigned as benign because they are not associated with a disorder, yet carrier status could be clinically crucial such as predicting ASD recurrence or the likelihood of comorbidities. On the other hand, assigning pathogenicity to these variants or genes is also an inaccurate term. This complexity highlights a critical gap in the current genetic interpretation frameworks. As researchers further study complex disorders (e.g., ADHD, OCD, bipolar disorder), there will be a growing need to adapt or develop new guidelines to more accurately address the nuances of polygenic diseases.

## Methods

### Family-based search for MES genes in SPARK

For our initial family-based screening of MES genes, we harnessed a dataset comprising 10,908 families sourced from SPARK ^15^ iWES v1 (including sequencing waves WES1-WES4), in which both parents and at least one ASD offspring underwent sequencing via WES. All genotypes were annotated with Ensembl Variant Effect Predictor (VEP) ^40^. We excluded genotype calls that did not adhere to Mendelian inheritance patterns within the family (accounting for an average of 0.72% of variants), effectively eliminating *de novo* mutations. Furthermore, during the initial screening phase, we omitted 113 families in which a proband exhibited a *de novo* rare deleterious variant (a PTV or a missense variant with a PolyPhen-2 ^41^ score ≥0.5 or SIFT ^42^ score ≤ 0.05; gnomAD v3 ^16^ non-neuro allele frequency of <1%) in the genes *CHD8, SCN2A, SYNGAP1, SHANK3,* or *ADNP*. These particular genes have a reported zero FDR for association with ASD from TADA ^4^, signifying a strong and monogenic-like association with ASD. Consequently, they were deemed unsuitable for the identification of MES genes. *PTEN* was not removed for the screen as deleterious variants occurred in controls.

We isolated ultra-rare deleterious variants in the families (a PTV or a missense variant with a PolyPhen-2 score ≥0.5 or SIFT score ≤ 0.05; gnomAD v3 non-neuro allele frequency of <0.1%). Though many more missense pathogenicity estimators are available ^43^, we opted for ones that scored for protein function impact without emphasis on species conservation or gene constraint (e.g., MPC^44^). Per our hypothesis, deleterious variants in single MES genes do not result in a strong phenotype and may not show as robust evidence of negative selection. For the same reason, we did not exclude genes based on LOEUF or pLI score. Additionally, we only included heterozygous genotypes on the reasoning that (1) homozygous combinations of ultra-rare deleterious variants are effectively unobserved, and (2) individuals homozygous for deleterious variants in a MES gene may have a separate non-ASD phenotype.

We first identified familial structures characterized by the inheritance of a specific ultra-rare deleterious variant (denoted as ‘A*’) by all offspring with ASD within the family, with ‘A*’ also being present in one non-ASD parent. Subsequently, we identified mirrored patterns of inheritance, focusing on a second variant in a distinct gene (’B*’). In such scenarios, ‘B*’ must derive from the parent who did not transmit ‘A*’. ‘A*B*’ must be present in all offspring with ASD and never in non-ASD siblings. Importantly, our methodology accommodated the potential existence of multiple ‘A*’ or ‘B*’ variants within the same gene in a single family. We quantified these patterns based on the number of unique families, rather than unique sites.

We next assessed the likelihood of observing ‘B*’ inheritance patterns within families exhibiting ‘A*’ inheritance. For this, we conducted binomial samplings to estimate the number of times we should observe any number of secondary independent gene hits:

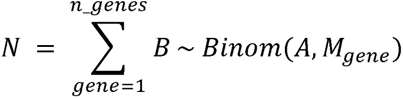

where B is the number of families with ‘B*’ inheritance, A is the number of families with ‘A*’ inheritance, and M_gene_ is the frequency of ultra-rare deleterious variants in a gene among the 21,570 non-ASD parents. This outputs N, the expected number of genes with B secondary hits given A.

Drawing from the frequency of ultra-rare deleterious variants per gene, calculated using parental data, our aim was to standardize N, representing the expected number of genes with secondary B hits, given A. Notably, larger genes, containing a higher occurrence of ultra-rare deleterious variants, might bear reduced significance when compared to smaller genes with fewer such variants. In our approach, for each pair of candidate MES genes, we multiplied N by the relative frequency of ultra-rare deleterious variants in the respective genes, designated as ‘N_norm’.

The gene possessing the highest number of ultra-rare deleterious variants, *TTN*, would have an adjustment of one with all other genes adjusted by factors less than one, contingent on their relative frequency of ultra-rare deleterious variants to *TTN*. To illustrate, consider the case of *SNAP25* and *SLPI*. Among the families analyzed, five exhibited the ‘A*’ inheritance pattern for *SNAP25*, while 21 displayed the same for *SLPI*. Two families featured the mirrored ‘A*B*’ inheritance pattern for both genes. According to binomial sampling, the occurrence of ‘5-given-2’ is expected approximately 15 times, whereas ‘21-given-2’ should transpire approximately 196 times (before normalization). Due to their smaller gene sizes, *SNAP25* and *SLPI* correspondingly exhibited ‘N_norm’ values of 0.061 and 0.174, indicating that the co-occurrence of these two genes inherited together is relatively unexpected and thus included as a candidate pair for further investigation. We kept pairs where ‘N_norm’ was less than one for both configurations of gene pairs.

### Additional variant types and frequencies

In addition to PTVs and missense variants of ultra-rare or rarer frequency, we tested three other stringencies: PTVs and missense variants of rare (1%) or rarer frequency, only PTVs of ultra-rare (0.1%) and rarer frequency, and only PTVs of rare or rarer frequency. For PTVs and missense variants of rare or rarer frequency, we retained 2,507 pairs of genes (2,146 total unique genes) where the normalized expected frequency was less than one in both ‘A*’ and ‘B*’ configurations. After filtering for non-ASD ‘A*B*’ in the SPARK v1 dataset, only 179 candidate pairs remained (339 total unique genes). For PTVs of rare or rarer frequency, we retained 834 (1,336 total unique genes) candidate pairs of which only 83 (161 unique genes) remained after SPARK filtering. For PTVs of ultra-rare or rarer frequency, we retained 692 candidate pairs (1,177 total unique genes) of which only 63 (122 unique genes) remained after SPARK filtering.

### Non-neuro cohorts

We included variants from the 3,202 individuals from the 1000 genomes project (1kGP)^17,18^. SIFT scores, PolyPhen-2 scores, and allele frequencies were extracted from GnomAD (v3.1.2)^45^.

Analyses for All of Us were performed in the workspace titled “Detecting the prevalence of ultra-rare gene mutations.” We selected samples with short read whole genome sequencing and excluded individuals with a survey response diagnosis of ASD (diagnosed with, receiving treatment for, or seeking treatment for ASD) or any disorder characterized under ‘mental disorder.’ The final cohort included 156,550 individuals of which 50.5% were of a self-reported non-white race and 57.0% were assigned female sex at birth. Due to the security constraints of the All of Us workbench, VEP was performed with SNPeff ^42^ (4.3t) as opposed to Ensembl. GnomAD v3 frequencies, Polyphen-2 scores, and SIFT scores were acquired from dbNSFP(v4)^47^. 98.8% of missense variants had an available Polyphen-2 and SIFT score. For high impact insertions and deletions that were not present in dbNSFP, we used the allele frequency in the 156,550 individuals as a proxy. To avoid disseminating individual-level data, we did not report the precise number of individuals with ‘A*B*’ for any gene pair (unless zero or greater than 20). For any single candidate gene, ultra-rare deleterious variants were always present in at least 20 individuals.

We additionally included individuals from the BioMe Biobank maintained by the Mount Sinai Health System. We isolated 16,586 individuals with genotype information derived from WES and excluded any individuals with a diagnosed or self-reported mental, behavioral, or neurodevelopmental disorders (ICD codes F01-F99). Of the final cohort, the mean age was 61.7 years, 56.2% were of self-reported male sex, and 56.9% were of self-reported non-white race. For consistency, VEP was performed in an identical manner as for the All of Us cohort.

### Equilibrium test and Bayes factors

We determined the frequency of ultra-rare deleterious variants per gene across all samples, encompassing both ASD and non-ASD individuals. The probability of ‘A*B*’ was computed as the product of the individual gene frequencies. Using the R function ‘pbinom,’ we designated this product as the probability of success for ‘A*B*.’ Subsequently, we calculated (1) the probability of observing the given number or a greater number of ASD individuals with ‘A*B*’, considering the total number of individuals with ASD, and (2) the probability of observing the given number or a lesser number of non-ASD individuals with ‘A*B*’ mutations, considering the total number of individuals without ASD. The equilibrium probability was then determined as the product of these two probabilities.

For the Bayes factor, the null hypothesis was the equilibrium probability, and the alternate hypothesis was the probability of observing the given number or a greater number of individuals regardless of ASD status with ‘A*B*’, considering the total number of individuals. The Bayes factor was calculated as the ratio of alternate and null equilibrium probabilities.

### Validation analyses

We procured ultra-rare synonymous variants across all cohorts, excluding those with a functional impact (e.g., missense, or more significant effects) on another gene. Ultra-rare synonymous variants were, on average, 1.83 times more frequent per gene than ultra-rare deleterious variants, which underwent further filtration for deleteriousness via VEP. We determined the ratio of ultra-rare synonymous to ultra-rare deleterious variants across all protein-coding genes. As some genes presented markedly divergent ratios, we excluded those beyond the >95th or <5th percentile thresholds. This adjustment predominantly impacted smaller genes where variants were only observed in a few samples.

Using ultra-rare synonymous variants, the initial screening (steps 1-3 of **Fig 2**) identified 690 candidate gene pairs, significantly fewer than the 1,916 pairs identified with deleterious variants. Subsequent analysis incorporating the complete SPARK v1 cohort (step 4 of **Fig 2**) further narrowed the candidate pairs to 129 (258 unique genes).

In the SPARK v1 cohort, we noted outlier individuals with high counts of ultra-rare synonymous variants. On average, the occurrence of ultra-rare synonymous variants was less than one per individual per gene. Among 258 unique genes from 129 candidate pairs identified with synonymous variants, four samples exhibited over 100 variants in single genes, 50-fold higher than the mean (SP0122762, SP0379928, SP0149880, SP0064380; **Fig S3D**). The excess synonymous variants in the four samples were unique and not cataloged in gnomAD v4, casting doubts on their quality. Contrastingly, such an excess was not observed with ultra-rare deleterious variants (**Fig S3D**). We subsequently removed these four samples from analyses.

For additional validation, we incorporated novel samples from SPARK iWES v2 (WES5). Quality control was identical to that of SPARK v1.

### Gene expression

We used nTPM values derived from RNAseq from the Human Protein Atlas (v23)^22^. The 50 tissue consensus used transcriptomics from both the Human Protein Atlas and the GTEx consortium ^48^. Expression for the 193 brain subregions was also from the Human Protein Atlas. To test if the 97 predicted MES genes were significantly enriched in a tissue or brain subregion, we sampled genes from all protein coding genes without replacement in 1000 iterations. For each iteration, we measured the fraction of sampled genes expressed (nTPM >1) in each tissue or brain subregion. We used the Shapiro–Wilk test^49^ (R command ‘shapiro.test’) to confirm sampled statistics were normally distributed (p>0.05). We then used the cumulative density function of the normal distribution to determine the significance of predicted MES gene enrichment per tissue or brain subregion.

Using the same two RNAseq datasets described above, we calculated the correlation of nTMP values across tissues and brain subregions. To assess the significance of MES pair co-expression, we calculated the co-expression correlation of all possible MES gene pairs and then calculated the p-value of real MES pairs using the R function ‘pnorm’. Hierarchical clustering of the co-expression of MES and other ASD genes was performed with the R function ‘hclust’ using the ‘ward.D2’ method and plotted with ‘ComplexHeatmap’^50^.

### Phenotypes of MES carriers

We utilized the Social Communication Questionnaire (SCQ) and basic medical questionnaire administered in the SPARK v1 cohort. For the SCQ total score, we removed 167 non-ASD siblings with a score at or above 13 (two standard deviations above the mean among non-ASD respondents). For the basic medical screening, we excluded questions related to birth defects (as they were uncommon) and environmental exposures (e.g., lead poisoning, fetal alcohol syndrome, traumatic brain injury).

## Acknowledgements

The authors wish to thank Dr. David Cutler for insightful feedback on the manuscript and Marina Natividad Avila for assistance with data access.

The All of Us Research Program is supported by the National Institutes of Health, Office of the Director: Regional Medical Centers: 1 OT2 OD026549; 1 OT2 OD026554; 1 OT2 OD026557; 1 OT2 OD026556; 1 OT2 OD026550; 1 OT2 OD 026552; 1 OT2 OD026553; 1 OT2 OD026548; 1 OT2 OD026551; 1 OT2 OD026555; IAA #: AOD 16037; Federally Qualified Health Centers: HHSN 263201600085U; Data and Research Center: 5 U2C OD023196; Biobank: 1 U24 OD023121; The Participant Center: U24 OD023176; Participant Technology Systems Center: 1 U24 OD023163; Communications and Engagement: 3 OT2 OD023205; 3 OT2 OD023206; and Community Partners: 1 OT2 OD025277; 3 OT2 OD025315; 1 OT2 OD025337; 1 OT2 OD025276. In addition, the All of Us Research Program would not be possible without the partnership of its participants.

The BioMe Biobank is supported by The Charles Bronfman Institute for Personalized Medicine at Mount Sinai. We want to further thank all participants and families that were a part of the research cohorts used in this research.

This study received funding from the Beatrice and Samuel A. Seaver Foundation (BM, JDB) and the National Institutes of Health (NIH), under grant numbers R01MH129724 (JDB, BM) and R01MH128813 (JDB).

## Author contributions

M.C. led the study design, conducted analyses, and authored the manuscript. B.M. contributed to the study design and manuscript editing and, along with J.D.M., oversaw project management. F.K.S. performed data generation and quality control related to the SPARK cohort.

## Data availability

Whole genome sequencing from 1kGP is freely available from The International Genome Sample Resource (https://www.internationalgenome.org/). All other individual-level data used in this study are not publicly available with access permission subject to review. The genetic and phenotypic data for SPARK can be requested at SFARI Base (https://www.sfari.org/resource/spark/). Access to the Sinai BioMe data can be requested through The Charles Bronfman Institute for Personalized Medicine (https://icahn.mssm.edu/research/ipm/programs/biome-biobank) and is subject to compliance with the Mount Sinai Health System’s data use agreements. All of Us controlled tier data is available for registered institutions and cohort selection is detailed in the workbench titled “Detecting the prevalence of ultra-rare gene mutations.”

## Code availability

Code and resources used in this study is available on GitHub at (https://github.com/MahjaniLab/MES_Code). For any further inquiries or requests for code not available in the repository, please contact the corresponding author.

