## Supplemental Figures for "Identification of moderate effect size genes in autism spectrum disorder through a novel gene pairing approach"

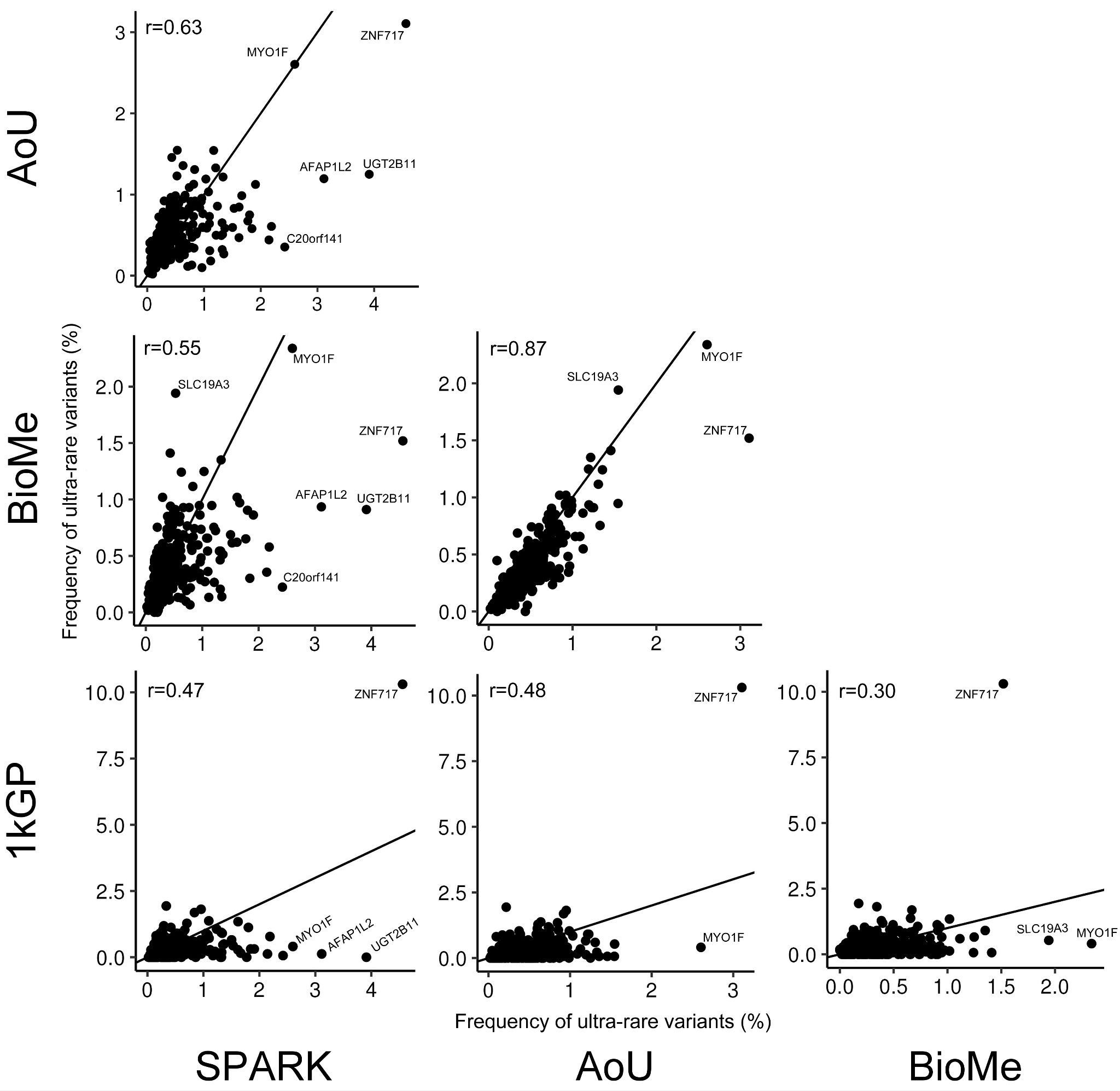

**Figure S1. Comparison of ultra-rare variant frequency by MES candidate gene across cohorts.** The percent of individuals per cohort with at least one ultra-rare deleterious variant in 318 candidate MES genes. The line represents linear regression. Pearson correlation coefficients are included for each plot.

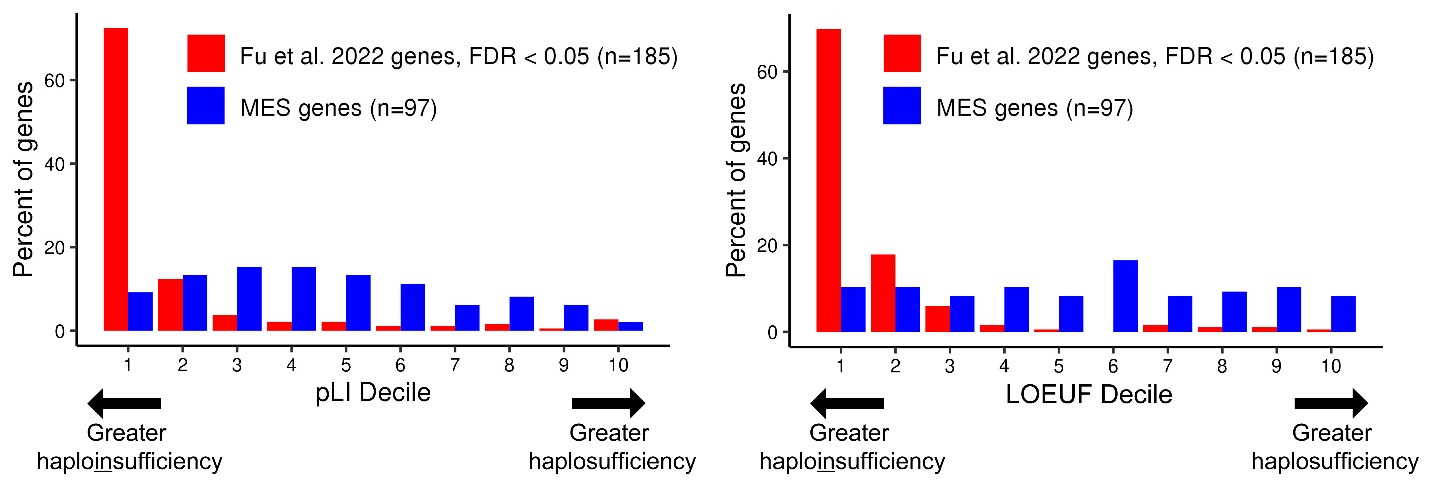

**Figure S2. Distribution of pLI and LOEUF scores for ASD genes.** Representation of MES genes and genes described in Fu et al. 2022 in decile bins created from the probability of loss of function intolerance (pLI) and loss-of-function observed over expected upper bound fraction (LOEUF) score of all protein coding genes.

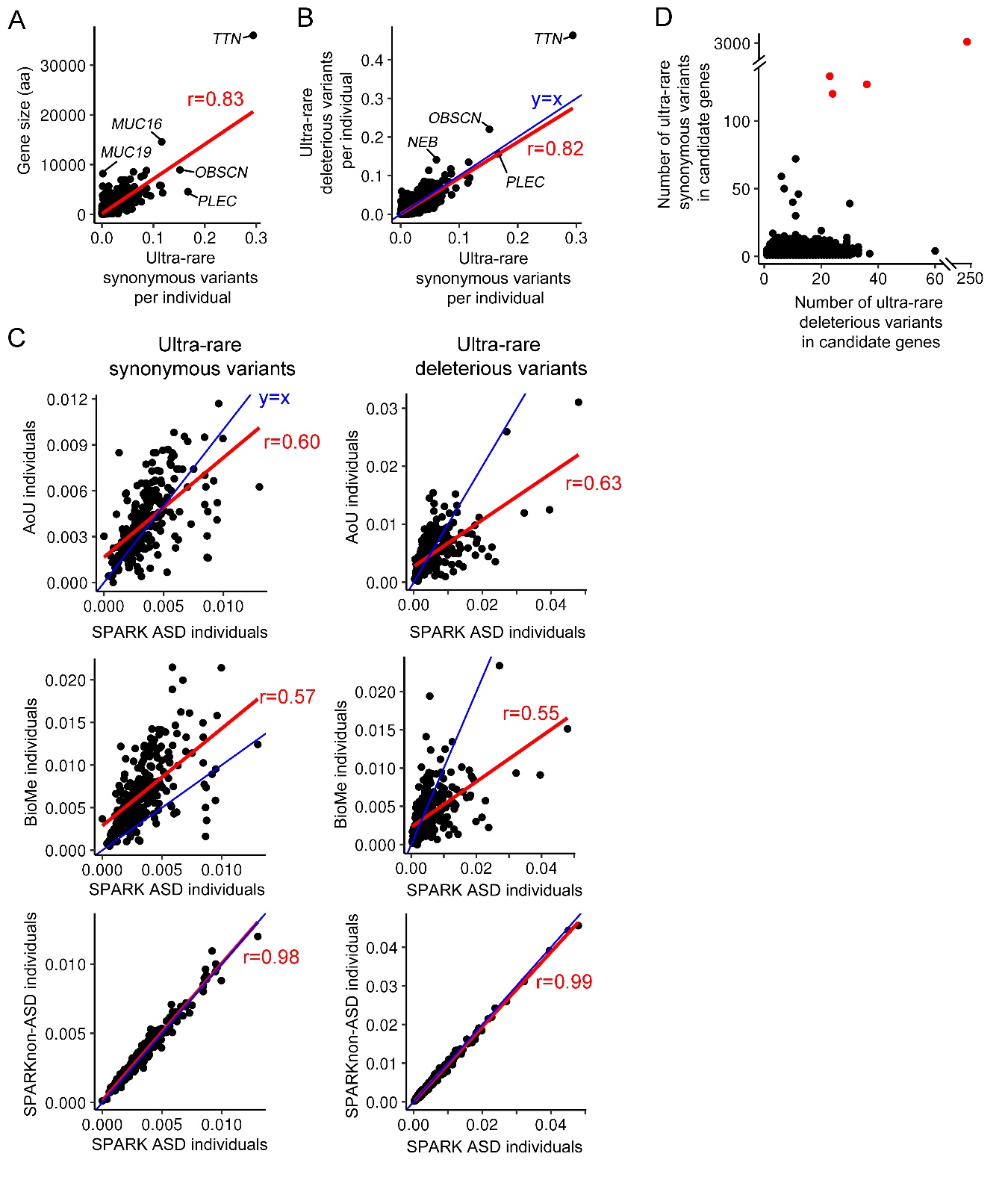

**Figure S3. Validation with ultra-rare synonymous variants.** (A) Mean number of ultra-rare (<0.1%) synonymous variants in SPARK parents versus gene size. The red line represents linear regression. (B) Mean number of ultra-rare synonymous variants in SPARK v1 parents versus the mean number of ultra-rare deleterious variants (missense and PTVs) as shown in Fig 1C. (C) Comparison of ultra-rare synonymous variant frequencies in candidate genes across controls cohorts to ASD individuals in SPARK v1. Of note, the candidate genes discovered with synonymous variants differed from those with deleterious variants. (D) The number of ultra-rare synonymous and deleterious variants in candidate genes among the SPARK v1 cohort. The four individuals highlighted in red were removed from analyses.

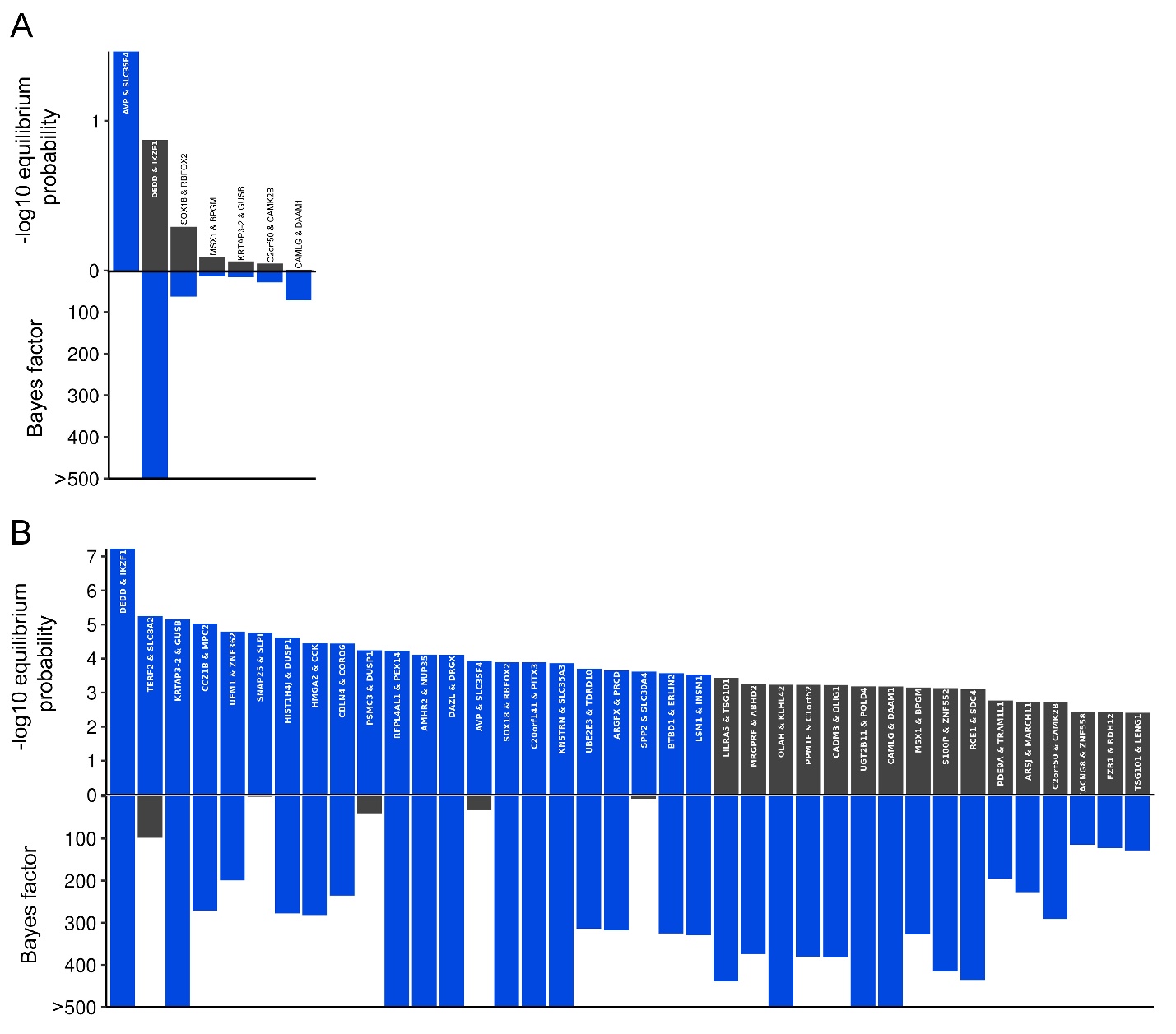

**Figure S4. Validation of MES genes with SPARK v2.** (A) As in Fig 1D, equilibrium probability and Bayes factors for the predicted MES gene pairs in ASD using SPARK v2 in place of SPARK v1. Bars in blue denote an equilibrium probability <0.05 or a Bayes factor >10. Only seven of the eight pairs with at least one A*B* in ASD are tested. (B) As in Fig 1D combining SPARK v1 and v2 samples.

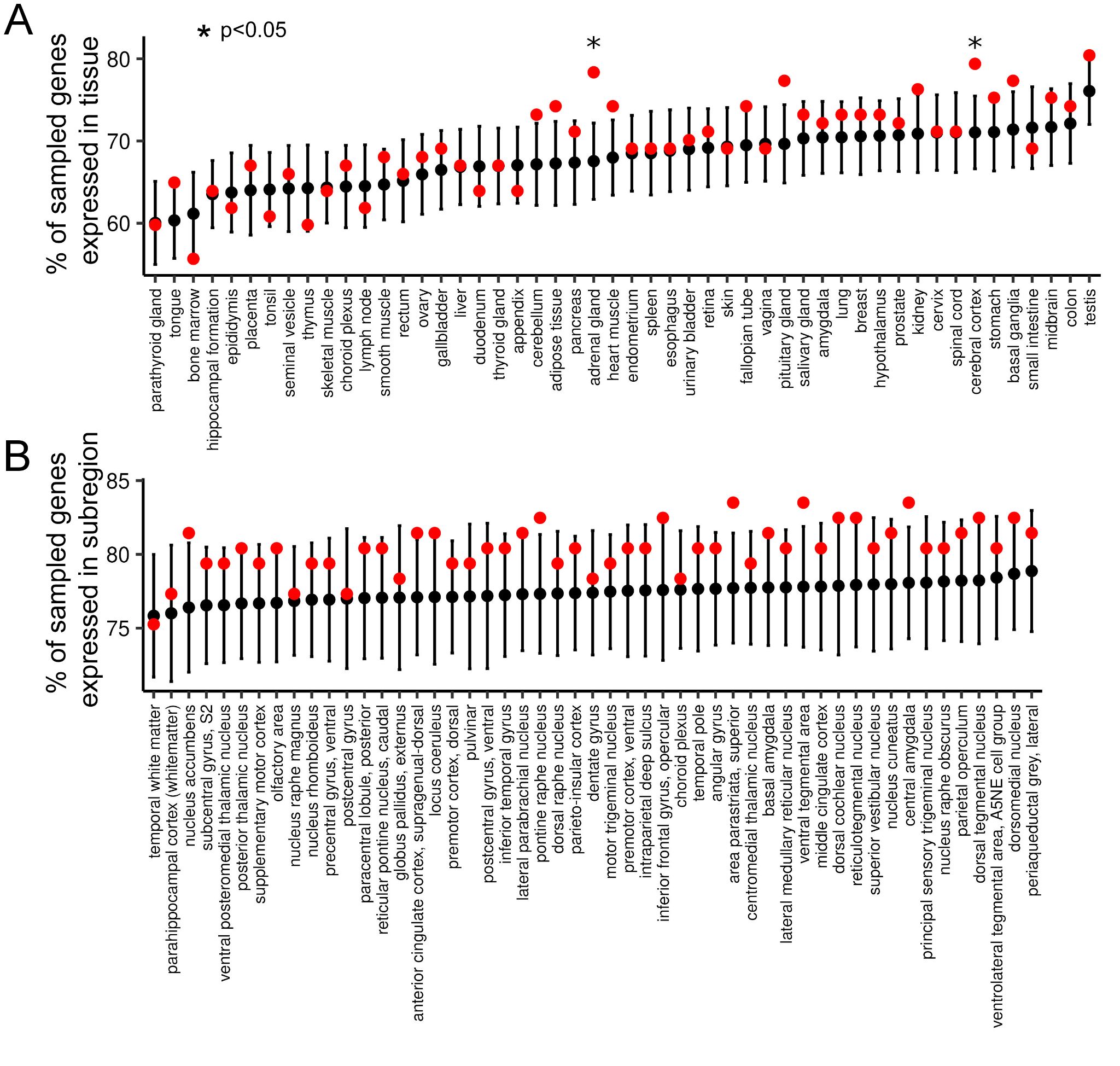

**Figure S5. Fraction of MES genes expressed per tissue compared to simulations.** (A) Red point: The actual proportion of the 97 predicted MES genes expressed (nTPM > 1) in the corresponding tissue. Black point: The average proportion of sampled protein-coding genes expressed in the respective tissue over 1000 iterations. Error bars denote the standard deviation of the samplings. Significant tissues are highlighted. (B) As in panel A for brain subregions. Only a subset of the 196 subregions is displayed.

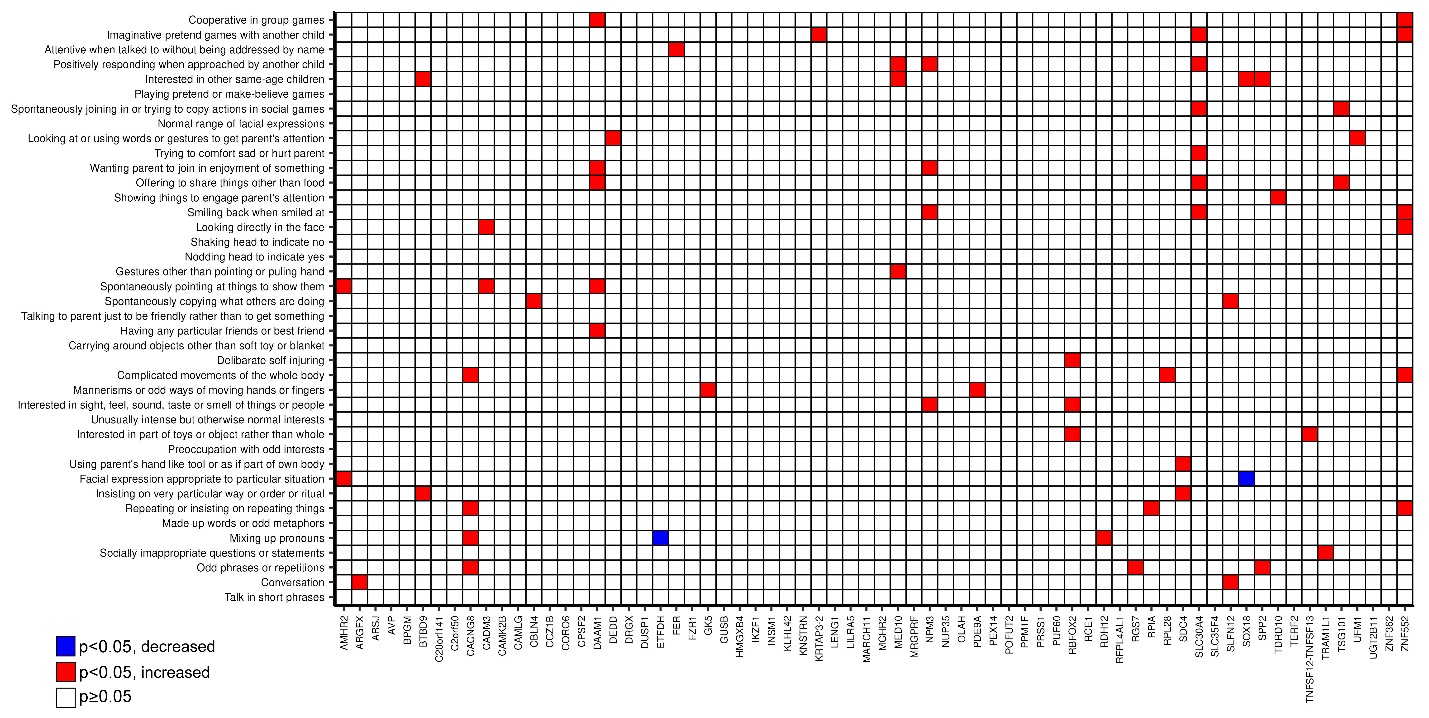

**Figure S6. Significant SCQ response skew in non-ASD carriers of MES genes.** Shown are the 68 MES genes with at least five non-ASD gene carriers with SCQ responses. Significant differences in carrier versus non-carrier responses are highlighted. In 63 of the 65 significant questions, carriers showed greater ASD-like responses, colored red. The remaining two showed fewer ASD-like responses, colored in blue.

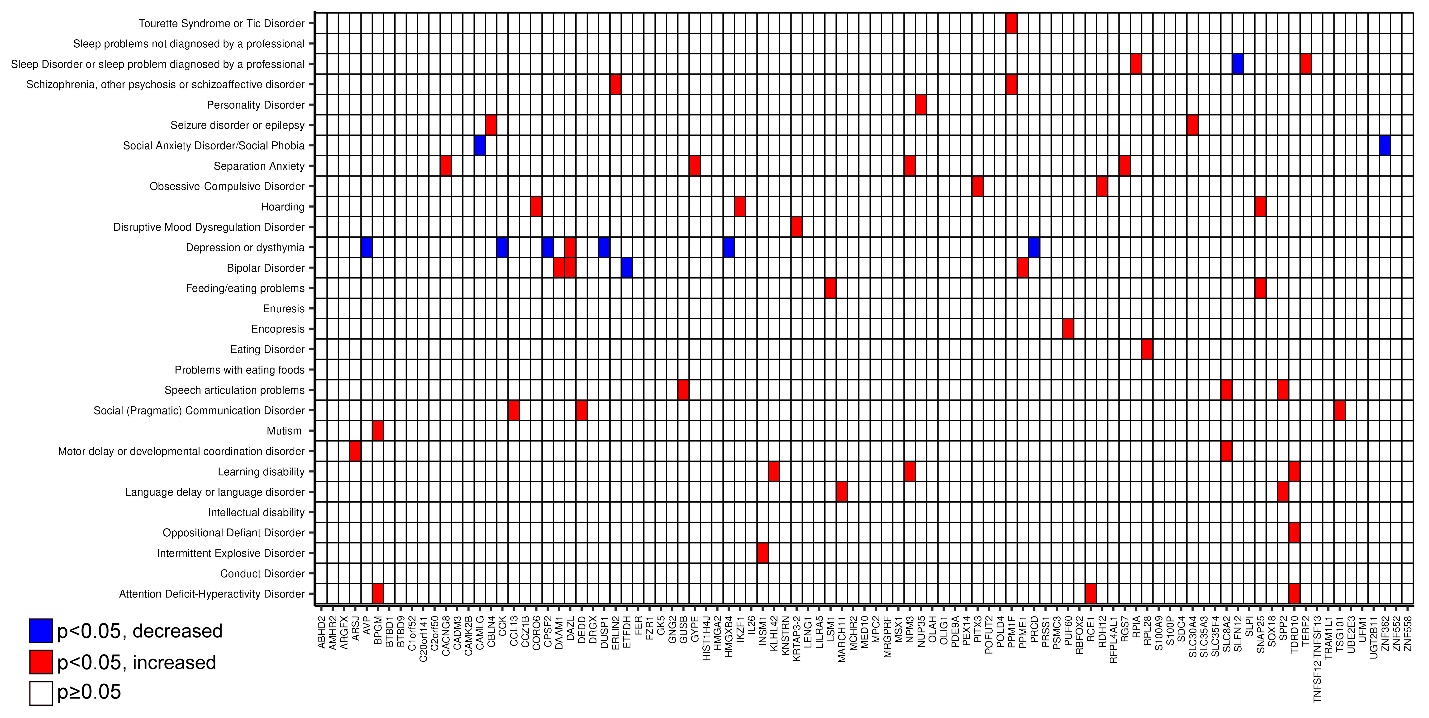

**Figure S7. Significant basic medical screening response skew in non-ASD carriers of MES genes.** Significant differences in carrier versus non-carrier condition frequencies are highlighted. In 51 of the 61 highlighted cases, carriers showed greater condition frequency, colored red. The remaining ten show decreased condition frequency, colored blue.

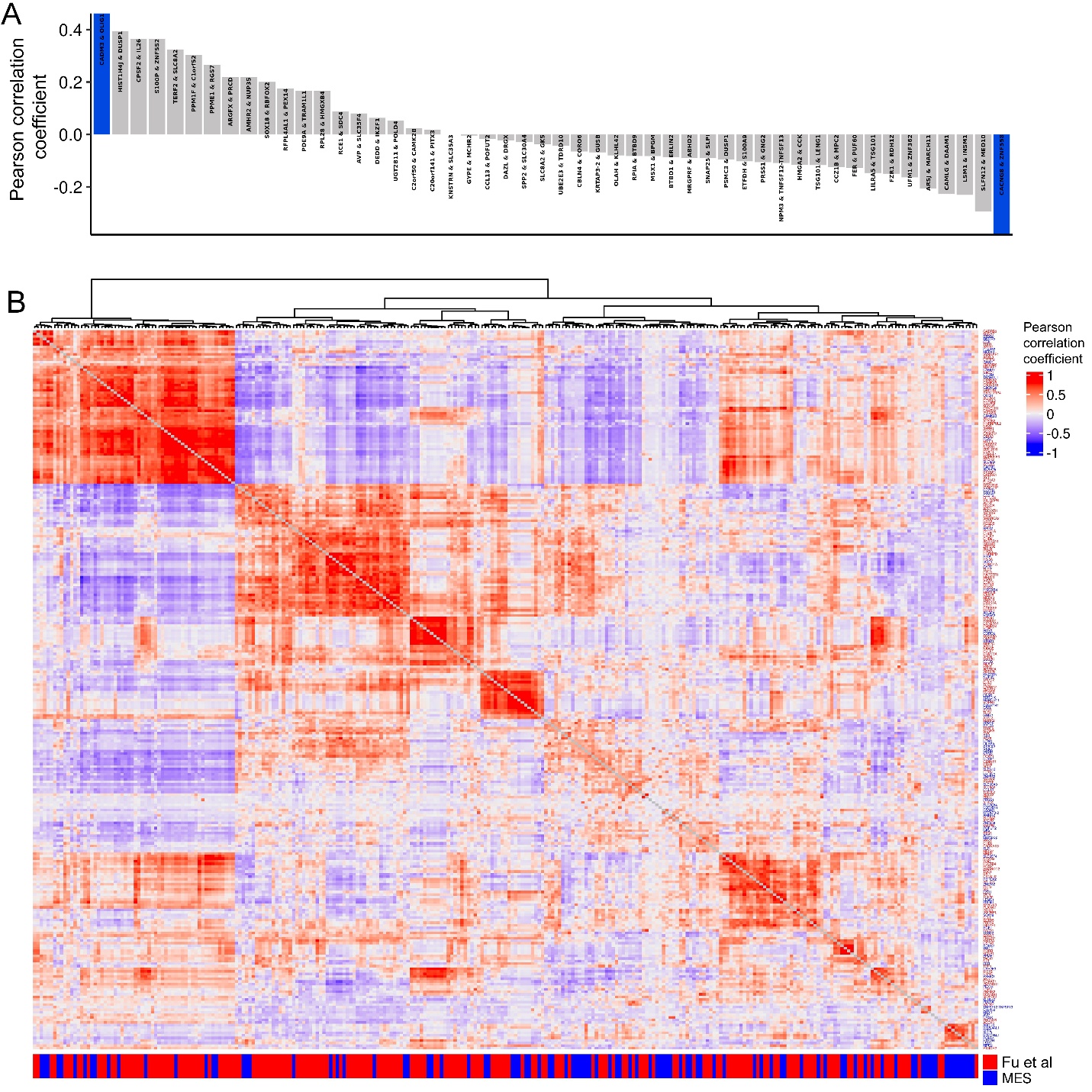

**Figure S8. Co-expression of ASD genes across 50 tissues.** (A) The correlation of nTMP values of paired MES genes. Bars highlighted in blue indicate significantly higher or lower correlation (p<0.05) compared to any two MES genes. (B) Clustered co-expression of the 97 MES genes (labeled in blue) and 185 genes ASD-associated genes described in Fu et al. 2022 (FDR < 0.05) (labeled in red).

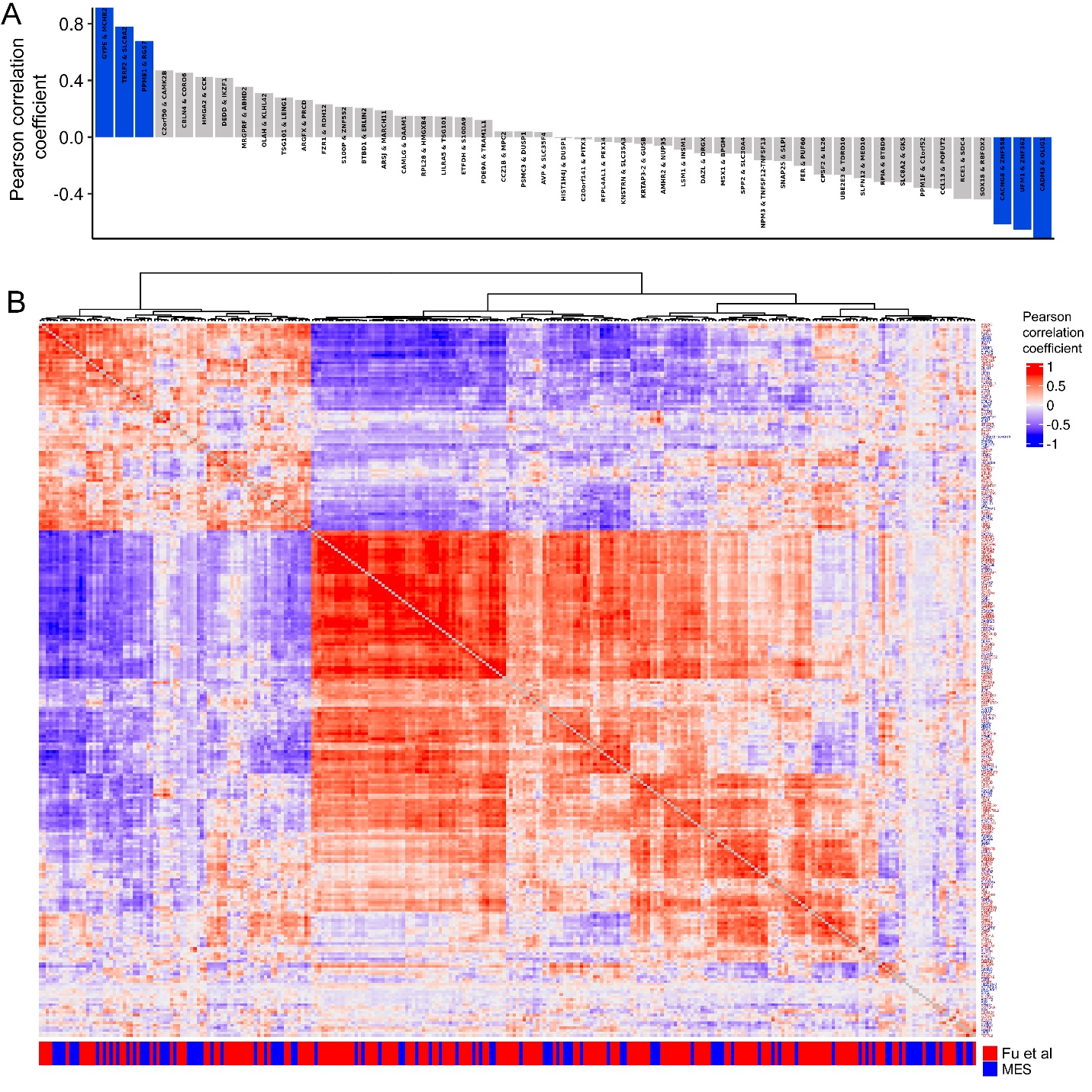

**Figure S9. Co-expression of ASD genes across 193 brain subregions.** (A) As in Fig S8, the correlation of nTMP values of paired MES genes. Bars highlighted in blue indicate significantly higher or lower correlation (p<0.05) compared to any two MES genes. Two pairs (*PRSS1/GNG2* and *UGT2B11/POLD4*) could not be tested due to undefined subregion expression. (B) As in Fig S8, clustered co-expression of the 97 MES genes (labeled in blue) and 185 genes ASD-associated genes described in Fu et al. 2022 (FDR < 0.05) (labeled in red).
